# Explant analysis of Utah electrode arrays implanted in human cortex for brain-computer-interfaces

**DOI:** 10.1101/2021.08.28.21262765

**Authors:** Kevin Woeppel, Christopher Hughes, Angelica J. Herrera, James Eles, Elizabeth C. Tyler-Kabara, Robert A. Gaunt, Jennifer L. Collinger, Xinyan Tracy Cui

## Abstract

Brain-computer interfaces are being developed to restore movement for people living with paralysis due to injury or disease. Although the therapeutic potential is great, long-term stability of the interface is critical for widespread clinical implementation. While many factors can affect recording and stimulation performance including electrode material stability and host tissue reaction, these factors have not been investigated in human implants. In this clinical study, we sought to characterize the material integrity and biological tissue encapsulation via explant analysis in an effort to identify factors that influence electrophysiological performance.

We examined a total of six Utah arrays explanted from two human participants involved in intracortical BCI studies. Two Pt arrays were implanted for 980 days in one participant (P1) and two Pt and two iridium oxide (IrOx) arrays were implanted for 182 days in the second participant (P2). We observed that the recording quality followed a similar trend in all 6 arrays with an initial increase in peak-to-peak voltage during the first 30-40 days and gradual decline thereafter in P1.

Using optical and two-photon microscopy (TPM) we observed a higher degree of tissue encapsulation on both arrays implanted for longer durations in participant P1. We then used scanning electron microscopy and energy dispersive X-ray spectroscopy to assess material degradation. All measures of material degradation for the Pt arrays were found to be more prominent in the participant with a longer implantation time. Two IrOx arrays were subjected to brief survey stimulations, and one of these arrays showed loss of iridium from majority of the stimulated sites. Recording performance appeared to be unaffected by this loss of iridium, suggesting that the adhesion of IrOx coating may have been compromised by the stimulation, but the metal layer did not detach until or after array removal.

In summary, both tissue encapsulation and material degradation were more pronounced in the arrays that were implanted for a longer duration. Additionally, these arrays also had lower signal amplitude and impedance. New biomaterial strategies that minimize fibrotic encapsulation and enhance material stability should be developed to achieve high quality recording and stimulation for longer implantation periods.

## Introduction

Intracortical brain-computer interfaces (BCIs) can restore function for people affected by significant paralysis by allowing the user to control an effector or assistive device with signals recorded in the brain. In recent years intracortical implants in motor cortex have been used for BCI control in primates and human participants with up to 10 degrees of freedom (Ajiboye et al. 2017; Bouton et al. 2016; Collinger et al. 2013; Hochberg et al. 2006; Santhanam et al. 2006; Velliste et al. 2008; Wodlinger et al. 2014). More recently, somatosensory feedback has also been added to these systems by stimulating through electrodes in the somatosensory cortex (Armenta Salas et al. 2018; Fifer et al. 2020; Flesher et al. 2016; Flesher et al. 2019; Flesher et al. 2021; Hughes et al. 2020; Hughes et al. 2020). Given that intracortical BCIs require surgical implantation, they must be stable over many years to be clinically viable. This issue has been studied in both humans and primates, demonstrating that signals can be reliably recorded from electrodes in the motor cortex for over 6 years when devices do not fail, although there is considerable inter-subject variability and signals typically deteriorate over time (Bullard et al. 2020; Chestek et al. 2011; Downey et al. 2018; Hughes et al. 2020; James et al. 2013; Simeral et al. 2011; Suner et al. 2005).

Changes in recorded activity can be caused by many factors including movements of the electrodes relative to the brain, encapsulation of the electrode sites, as well as material degradation and failure (Kozai et al. 2015; Prasad et al. 2014; Woeppel et al. 2017). These factors can be broadly grouped into multiple failure categories, including material and biological failure (James et al. 2013).

Biological failures can occur as a result of the host tissue reactions to the implant. The traumatic nature of the implant leads to glial activation and encapsulation of the implant in a glial sheath (Polikov et al. 2005; Salatino et al. 2017). The glial sheath creates a physical barrier between the electrode and the neurons, while the extensive inflammation damages healthy neurons and may cause a neuron dead zone around the implant (Buzsáki 2004; Schwartz et al. 2006). One recent study examining brain tissue from a human patient implanted with a Utah microelectrode array for seven months found a substantial degree of tissue damage which correlated with decreased recording performance.(Szymanski et al. 2021) In addition to central nervous system (CNS) reactions, the meninges can grow under the electrode. Meningeal encapsulation is highly collagenous and originates from non-CNS tissues. Substantial undergrowth of meningeal tissues can result in displacement of the electrode sites or complete ejection of the device from the CNS. (Woolley et al. 2013) Subsequent device ejection is the most prevalent cause of chronic device failure in non-human primates, accounting for nearly 30% of chronic failure (Barrese et al. 2016; Dunlap et al. 2020) Longer experimental times increase the chance of meningeal undergrowth and eventual ejection of the recording device from the host tissues (Barrese et al. 2016; Degenhart et al. 2016; Rousche and Normann 1998).

Material failures include metal corrosion, insulation cracking, and insulation delamination. These material failure modes often increase in likelihood as time progresses. The parylene-C insulation commonly used for Utah style intracortical arrays can crack and delaminate, shunting current to the biological tissues (Caldwell et al. 2020; Prasad et al. 2014; Schmidt et al. 1988; Xie et al. 2014). The metal tips of Utah arrays, most commonly platinum or iridium oxide, are generally stable *in vitro*, but may be eroded away by aggressive stimulation (Negi et al. 2010) or the comparatively harsh *in vivo* environment (Negi et al. 2010). Furthermore, use of the electrodes for stimulation can impact the rate of tip degradation (Cogan 2008; Gilgunn et al. 2013).

To establish stimulation limits for these clinical studies, experiments were performed in non-human primates and showed that frequent microstimulation over six months did not cause more loss of neurons around the electrode tips than insertion of the devices themselves and that stimulation had no behavioral effect for tasks that required tactile feedback (Chen et al. 2014; Kim et al. 2015). Using these established parameters, we would not expect stimulation to cause further damage to the brain tissue after implantation or have deleterious effects on behavior. In fact, stimulation over five years in a participant with these established parameters has not resulted in significant differences in signal between stimulated and non-stimulated arrays and detection thresholds have improved over time (Hughes et al. 2020). However, to our knowledge there have been no post-implant examinations of the material properties of intracortical arrays implanted in humans. Here we examine the extent to which any material degradation occurred on explanted human intracortical electrodes, which will aid in the design and development of robust BCIs for long-term clinical use.

In this work, electrodes explanted from two human participants were examined to determine the extent of tissue encapsulation and material failure and to assess how these factors affected chronic recording performance. These electrodes were implanted for different lengths of time and were surgically explanted: 987 days for the two arrays in participant 1 (P1) and 182 days for the four arrays in participant 2 (P2). Both arrays in P1 and two of the arrays in the P2 had platinum tips and were used for recording only, while the other two of the arrays in P2 had sputtered iridium oxide (IrOx) tips and were used for both stimulating and recording (Negi et al. 2010). First, the extent and nature of the tissue encapsulation of the arrays was investigated using optical microscopy and two-photon microscopy (TPM). Following this, the electrode arrays were examined with scanning electron microscopy (SEM) and energy-dispersive x-ray spectroscopy (EDS) to evaluate the extent of material damage. Finally, we compared the results of these analyses to endpoint recording performance of the devices and characterized the relationship between electrical stimulation and material degradation.

## 1. Methods

### 1.1 Participants

These studies (NCT01894802 and NCT01364480) were conducted under Investigational Device Exemptions from the U.S. Food and Drug administration and were approved by the Institutional Review Boards at the University of Pittsburgh (Pittsburgh, PA) and the Space and Naval Warfare Systems Center Pacific (San Diego, CA). Informed consent was obtained before any study procedures were conducted. Two participants were implanted with microelectrode arrays in the brain. The first subject (P1) was implanted with two intracortical Pt microelectrode arrays (4 mm × 4 mm, Blackrock Microsystems, Salt Lake City, UT, USA) each with 96 wired electrode shanks (length 1.5 mm) in a 10×10 grid in the participant’s left motor cortex (Figure 1). The second subject (P2) was implanted with two Pt microelectrode arrays (Blackrock Microsystems, Salt Lake City, UT) in the left somatosensory cortex and two iridium oxide (IrOx) microelectrode arrays in the left posterior parietal cortex. Each Pt array in the somatosensory cortex consisted of 88 wired electrodes in a 10×10 grid while each IrOx array in the posterior parietal cortex consisted of 32 wired electrodes distributed throughout a 6×10 grid (Figure 1). Following implantation of the arrays into P2, it was discovered that the implant locations were posterior to the intended sites. Following which, the pedestals were removed, and a second implantation was performed two months later.

**Figure 1.**
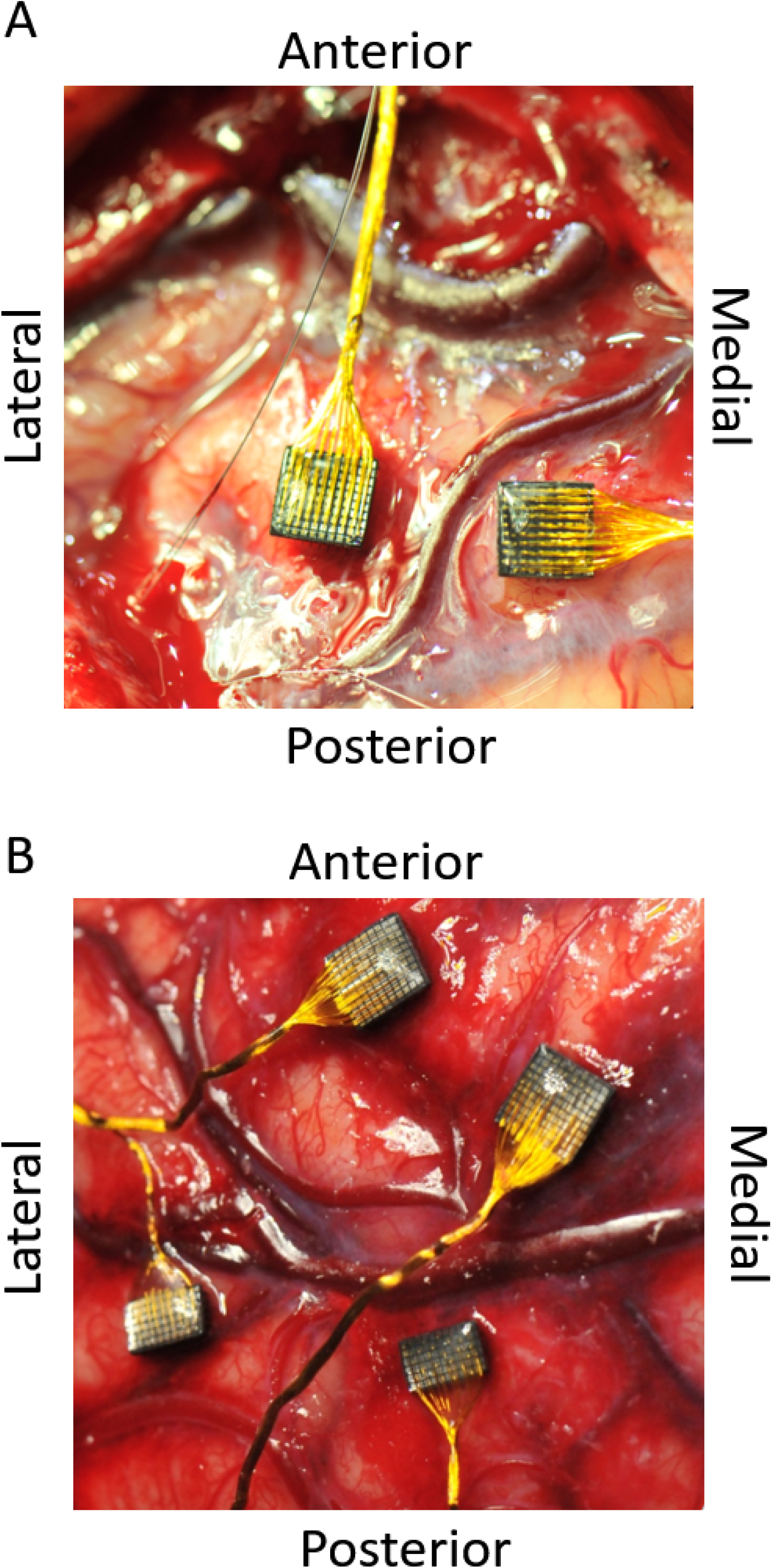
Six electrode arrays were implanted in two participants: two recording arrays in P1 motor cortex, two recording arrays in P2 somatosensory cortex, and two stimulating arrays in P2 medial parietal cortex. Intraoperative images of implanted arrays in P1 (A) and P2 (B).

### 1.2 Neural recording and signal quality metrics

Neural data were collected for both P1 and P2 using Neuroport Neural Signal Processors (Blackrock Microsystems, Salt Lake City, UT). At the beginning of each test session, a threshold for all channels was set at -5.25 (P1 before day 565) and -4.5 (all other test sessions) times the root-mean-square voltage. Data were collected across 287 sessions spanning 33 months for P1 and 40 sessions across four months for P2. No recordings were done for the final two months of P2’s implant as the percutaneous pedestal connectors had been removed to prepare for the reimplant.

One of the main goals of the clinical study was to provide the participants with high degree-of-freedom control of a robotic arm. To accomplish this, participants performed a brain-computer interface calibration paradigm at the beginning of a test session. We used three minutes of data collected during this calibration procedure to run spike sorting analyses offline. The sorting method, described in detail in Downey et al., 2018 (Downey et al. 2018) used principal component analysis (PCA) to separate units, defined as threshold crossings from an individual electrode, based on the similarity of their waveform shape. Characteristics for each unit were then calculated. Peak-to-peak voltage (Vpp) was defined as the voltage difference between the peak and the trough of the average waveform for each unit. Since there could be more than one unit identified per electrode, the unit with the maximum Vpp was chosen to represent the signal quality for the given electrode. Electrodes were considered to be viable if they contained waveforms with a minimum Vpp of 30 µV and a minimum firing rate of 0.25 Hz.

#### 1.2.1 Impedances

Electrode impedances were measured for both participants using the NeuroPort patient cable data acquisition system (Blackrock Microsystems, Salt Lake City, UT). For P1, impedances were measured at the beginning of a test session once a month. Impedances values for P2 were measured at the beginning of each test session. The system delivered a 1 kHz, 10 nA peak-to-peak sinusoidal current to each implanted electrode for one second.

#### 1.2.2 Intracortical stimulation and calculated metrics

Seven test sessions across approximately one month involved microstimulation on the IrOx arrays. Stimulation was delivered using a CereStim R96 multichannel microstimulation system (Blackrock Microsystems, Salt Lake City, UT). Pulse trains consisted of cathodal phase first, current-controlled, charge-balanced pulses delivered at frequencies from 20-300 Hz and at amplitudes from 1-100 μA. The cathodal phase was 200 μs long, the anodal phase was 400 μs long, and the anodal phase was set to half the amplitude of the cathodal phase. The phases were separated by a 100 μs interphase period. Stimulus pulse trains were varied in terms of amplitude, frequency, and train duration.

The voltage transients associated with each stimulus pulse were recorded using National Instruments data acquisition modules. Voltage traces were displayed in real time using LabView and saved to disk for analysis. Interphase voltage was measured as the voltage at the end of the interphase period immediately prior to the anodal phase for a given stimulation pulse. The total charge delivered to each electrode was calculated across all stimulation experiments using the charge delivered during the cathodal phase.

### 1.3 Explanted array handling before imaging

The two Pt arrays in P1 were explanted on day 987 post-implant and the four arrays in P2 were explanted on day 182. Following explantation, all arrays were removed from their wire bundles by clipping the wires proximal to the probe and were washed with saline. The P1 arrays were immediately fixed in formalin and then transferred to PBS bath for storage. Immunohistochemical staining procedure was performed on these two arrays with the goal of identifying neuron (NeuN) and microglia/macrophage (Iba-1). The staining process involves incubation of the arrays with primary antibodies solutions overnight, with secondary antibodies for 4 hours followed by Hoescht solution for 20 min for nuclei staining. The antibody staining was unsuccessful, and only nuclei stain was used for the tissue analysis. The P2 arrays were fixed 2 months post-implant, and one of the Pt arrays had visible tissue encapsulation and was imaged using TPM. Because these arrays were not immediately fixed, we did not perform immunostaining, and only characterized the collagen structure, which can be stable without the fixation.

After optical and TPM imaging, two arrays explanted from P1 were sent to the FDA for initial analysis. The arrays were initially imaged with an environmental SEM, then enzymatically cleaned with Asepti-Zyme neutral pH enzymatic instrument presoak/cleaner (4ml in 250ml saline) at 37°C for 90 minutes, followed by Getinge Clean Enzymatic detergent (1ml in 250ml saline) at 37°C for 90 minutes, and then by MetriZyme detergent (1ml in 250ml saline) 37°C for 90 minutes. Samples were then thoroughly washed with water and air dried, ready for SEM imaging. This process was effective at removing some of the tissue and revealing the electrode tip/shank for material analysis. Arrays from P2 did not undergo the enzymatic cleaning procedure. All arrays were stored adhered to copper tape, tips up.

### 1.4 Electrode Imaging

Explanted electrodes were first characterized by optical and two-photon microscopy to assess the degree of tissue encapsulation. For TPM, we used a two-photon laser scanning microscope with a Bruker scan head (Prairie Technologies, Madison, WI), TI:sapphire laser tuned to 920 nm (Mai Tai DS; Spectra-Physics, Menlo Park, CA), light collection through non-descanned photomultiplier tubes (Hamamatsu Photonics KK, Hamamatsu, Shizuoka, Japan), and a 10x or 16x, 0.8 numerical aperture water immersion objective (Nikon Inc., Milville, NY). Laser power was maintained between 20-40 mW. For each electrode tip, Z-stacks were collected with filters to resolve second harmonic generation (SHG) at half the laser wavelength (∼460nm), which enabled intrinsic imaging of collagen-I representing the meningeal encapsulation. Images along the length of the electrode shanks were collected as Z-stacks. Z-stack images were either collected at specific regions of interest, or in a grid at all locations across the face of the electrode array. Grid images were automated by the Prairie software with a 10% overlap between images. All image stitching and subsequent image processing was conducted with ImageJ software (NIH). Electrode integrity was characterized by scanning electron microscopy (SEM) and energy-dispersive x-ray spectroscopy (EDS). Samples were washed, dried under alcohol, and sputter-coated with 4nm Au/Pd. Images were taken by JSM 6335F electron microscope. EDS was taken by Zeiss Sigma 500VP, excluding Au and Pd from quantification.

Using the SEM and optical images, a qualitative category of ‘non-degraded/unencapsulated’ or ‘degraded/encapsulated’ was assigned to each electrode based on the degree of damage to the tip or shank, or the level of encapsulation around the electrode (**Figure S1**). Arrays explanted from P1 were more extensively cleaned prior to imaging, and the encapsulation score was based on optical images of the explanted arrays. Encapsulation on arrays from P2 was determined by examining the SEM images. Degraded electrode tips were defined as having obvious and substantial surface defects in the metal coating, including pitting of the metal, flaking of the metal, and exposure of the underlying silicon. Degraded shanks were defined relative to the parylene insulation, with defects including insulation cracking along the shank, peeling of the insulation away from the shank near the tip, and other obvious defects in or below the insulation. These categories were compared to EDS images, confirming the presence/absence of metalation at the tip (Pt/IrOx). Electrodes which could not be quantified, due to breakage during removal or gross encapsulation, were assigned a null score and excluded from analysis.

### 1.5 Statistics

Changes in signal and impedances over time were assessed using linear regression. For impedances, data were log-transformed because data did not follow a linear trend. For P2 impedances and Vpp, we excluded data prior to day 30 for regression because the impedances measured in this range were highly variable.

Total charge delivered, minimum interphase voltages, and charge delivered after exceeding an interphase voltage of -0.6V were compared between the two electrode arrays that had received stimulation using Mann-Whitney tests. We used a non-parametric test because the data was determined to not be normally distributed using an Anderson-Darling test. We used a Fisher exact test to determine if there was a significant relationship between an electrode’s material properties (undamaged or damaged) and the length of implantation (Pt arrays in P1 vs. P2) or if it received stimulation (P2 IrOx arrays, yes or no). We further quantified if there was a relationship between both total charge injected and charge injected with interphase voltages below -0.6V on stimulated electrodes and their material properties (undamaged or damaged) using logistic regression. Electrode categories were compared to impedances and Vpp using Mann-Whitney tests. We used a non-parametric test because the variances between groups were not the same.

## 2 Results

### 2.1 Signal amplitude and impedances decreased over time

Changes in the impedances and peak-to-peak voltages over time were observed on implanted electrodes in both participants (Figure 2). Impedances decreased over time on electrodes implanted in P1 (p<0.001, log-transformed linear regression, Figure 2A). For P2, the starting impedances of IrOx electrodes were lower than the platinum electrodes, which is consistent with the manufacturer’s specification (Negi et al. 2010). From day 1 to 20 we observed an increase in impedances. The initial increase in impedance reversed after one month (30 days), and a significant downward trend in impedances was observed until the end of recording for both the IrOx (p<0.001, linear regression) and platinum arrays (p<0.001, linear regression). Impedances gathered from P1 eventually stabilized after approximately two years. The difference between the final impedance values recorded in P1 and P2 can be explained by the difference in length of implantation. Previous studies have determined that impedance values of stimulated and non-stimulated intracortical electrodes decrease dramatically over the first couple of years after implantation in humans (Hughes et al. 2020) and monkeys (Chestek et al. 2011; Suner et al. 2005). Since P2 was implanted for a significantly shorter period, we would expect the electrode impedance values to be larger and more variable, which the data supports.

**Figure 2:**
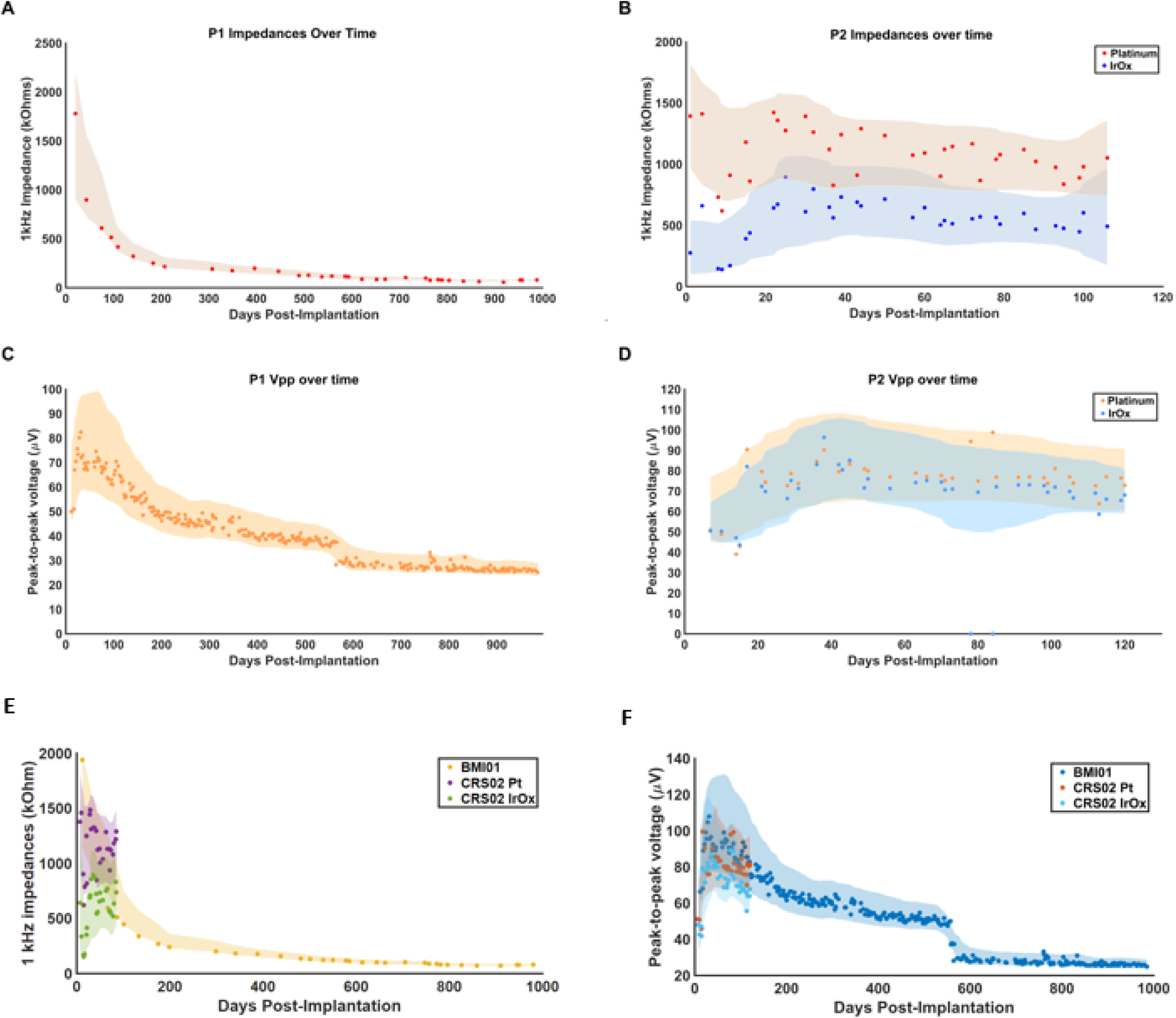
Impedances and peak-to-peak voltages decreased over time. Data points represent the median across electrodes for a given test date. The shaded regions show the interquartile ranges smoothed with a nine-point moving average filter with a triangular kernel. Median impedances recorded on (A) P1 electrodes and (B) P2 electrodes across the length of implant. Impedance measurements on P1 were not conducted with the same temporal resolution as P2. Different colors represent platinum or IrOx for P2 as indicated in the legend. Vpp recorded on (C) P1 electrodes and (D) P2 electrodes across the length of implant. For P1, there was a discontinuity in the Vpp at day post-implant 550 due to a change in the RMS threshold from -5.25 to -4.5. Overlayed impedances and Vpp for P1 and P2 are shown in (E) and (F), respectively.

In the same manner as the impedance measurements, an initial increase in Vpp was observed for both P1 and P2. However, after an increase in the first 30 days, the measured Vpp from P1 and P2 exhibited a downward trend (p<0.001, linear regression, Figure 2C,D). The rates of decrease in the Vpp between day 30 and 120 for P1 and P2 were -4.0 µV/month and -4.86 µV/month, respectively. Median Vpp decreased by 52% across 550 days in P1 and by 14% across 90 days in P2. The median Vpp for P1 leveled off at approximately 25-30 µV.

### 2.2 Encapsulating tissues were apparent on multiple arrays

Based on the gross optical micrographs, both P1 arrays had a significant degree of adherent tissue on the electrode base and shanks. For the P2 arrays, one of the Pt arrays and one of the IrOx arrays showed some tissue deposits while the other arrays were clean. The nature of the encapsulating tissue was examined with TPM, measuring the second harmonic signal characteristic of collagen. For the more heavily encapsulated P1 arrays, the encapsulation sheet was found both along the shanks of the array (Figure 3A-D) and at the base (Figure 3E-H (P1)). Strong second-harmonic signal within the tissue sheet confirmed that it was primarily composed of collagen-I fibers (Figure 3C,D,G,H). After further examining the indicated electrodes and staining for cell nuclei, we observed that the encapsulation was highly cellularized (Figure 3C,D,G,H). In addition, the encapsulation continued down the shank of the electrode, with cellular and collagenous material detected along the shanks and tips of the array. On the underside of the array at the base of the shanks the encapsulation was not homogenous, instead exhibiting greater second harmonic signals nearer to the edges (Figure 3G) while having greater cell density nearer the center (Figure 3H). SHG imaging is also a good tool for detecting blood vessels because of the strong presence of collagen in the vessel wall, however we did not observe clear blood vessel structure in the P1 explants.

**Figure 3:**
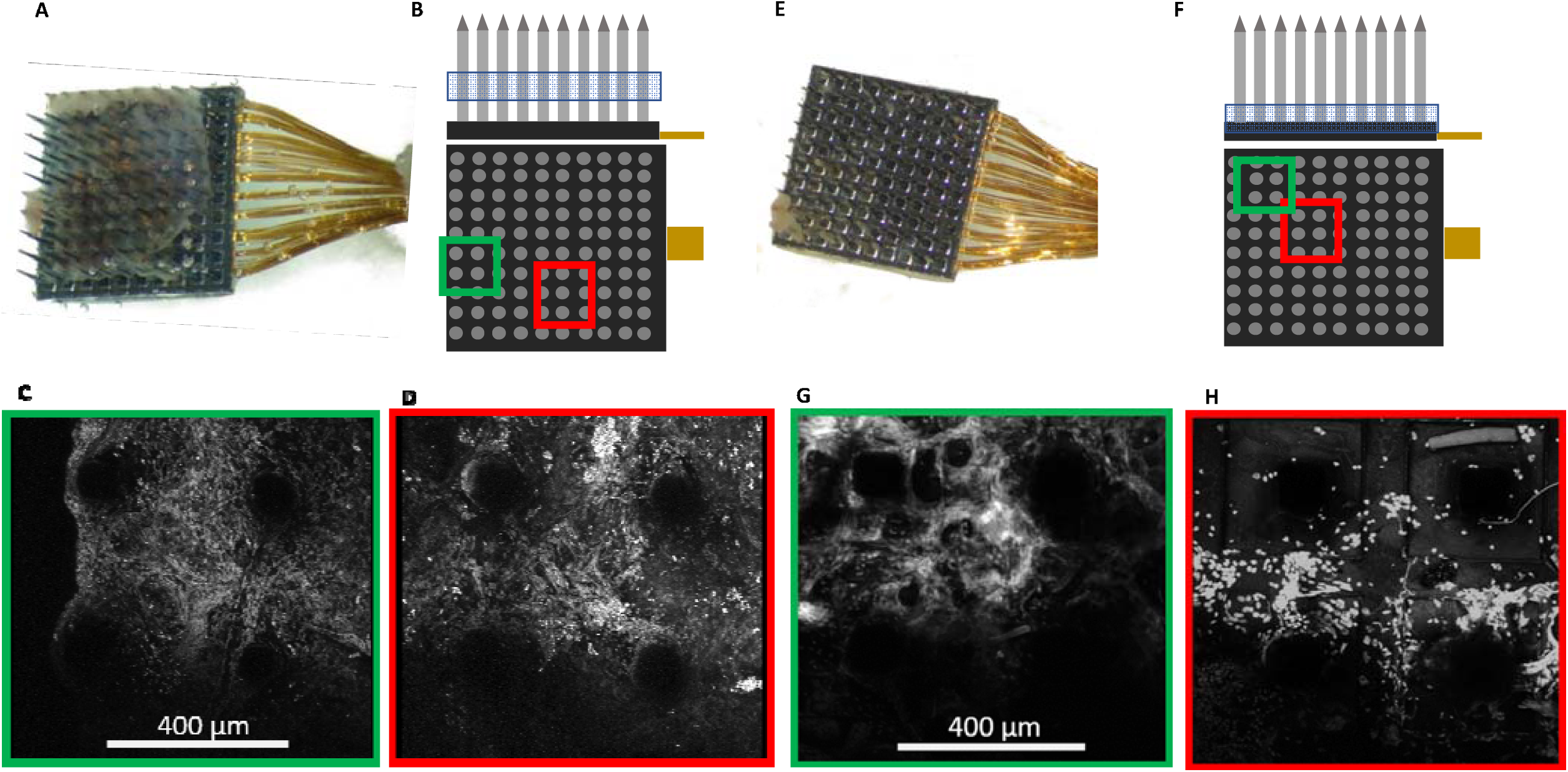
Characterization of the encapsulation of the electrodes. Arrays were imaged with an optical microscope in air. Both arrays are from P1. The encapsulation of array (A) was further examined with TPM. (B) The location of 2P imaging along the Z axis and select electrode shanks. The array was stained for cell nuclei and zoomed-in images were taken of the green (C) and red (D) regions. In both regions there is prominent second harmonic signal, indicating the presence of collagen. The array in (E) was chosen to display the lack of homogeneity of the encapsulating tissues. (F) Location along the z-axis (blue box) and 2 selected areas further imaged. 3D rotation images were generated displaying the tissue encapsulation and nuclei staining from the regions highlighted in green (G) or red (H). The outer image (G) displays high second harmonic signal while the inner image (H) has elevated cell counts, demonstrating the heterogeneity of the encapsulation.

For the posterior Pt array in P2, the encapsulation tissue covers the whole array (Figure 4B) and the TPM imaging from the side revealed significant tissue covering the majority of the electrode tips. Here, we identified clear vascular architecture in the encapsulation tissue (Figure 4D, highlighted in blue). The blood vessel in the encapsulation tissue was traced and super-imposed to the image of pial vasculature observed pre-implantation (Figure 4E). As can be seen in Figure 4F, the blood vessel traces match the pia vasculature. This indicates that the blood vessels observed to be at the tip of this array were pial blood vessels. Two mechanisms may lead to this: 1) the array did not fully penetrate the pia at the time of implantation; 2) the array was successfully implanted in the brain parenchyma and the pia membrane was pulled out with the array. Since we were able to obtain high quality single unit recordings from this array even from the affected region, the first potential mechanism was ruled out. Therefore, we conclude that at least some of the tissue on this explant is pia membrane that was pulled out with the device, not fibrotic scar tissue formed as the result of foreign body reaction.

**Figure 4:**
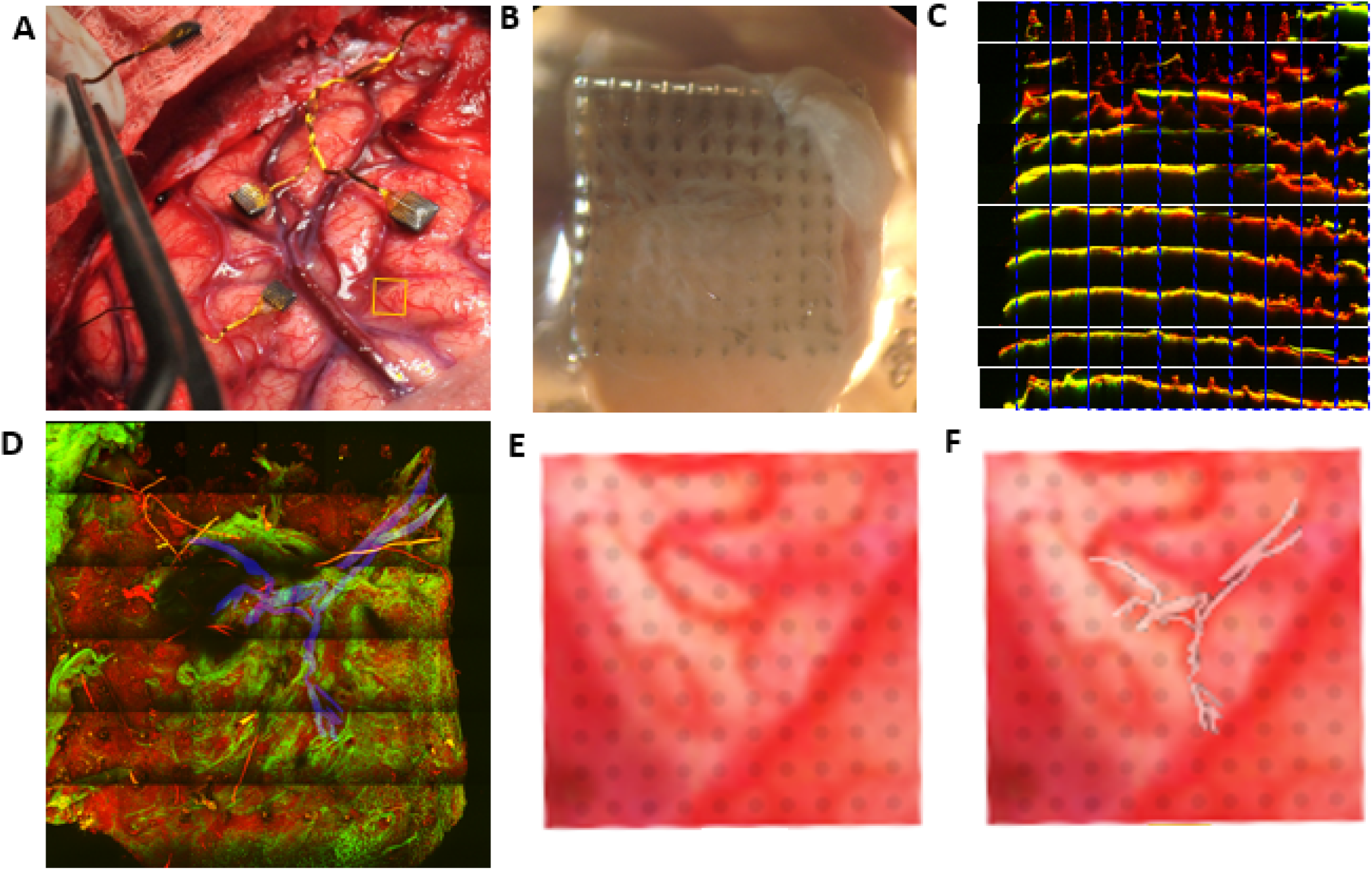
Brain vascularization can be visualized on one of the explanted arrays from P2. The pre-implant location is indicated with a yellow box (A). (B) Optical image of the array showing tissue coverage. (C) TPM of the shanks of the electrode, with green denoting second harmonic signal from collagen and red denoting the autofluorescence of the device. Each electrode was imaged and separated by row (side view). Most of the electrode tips are covered by collagenous tissue. (D) TPM image of the array looking from the tips downward, with a portion of vasculature marked in blue. (E) zoomed in image from (A) where electrode shanks are superimposed on the underlying vasculature. (F) The vasculature visualized in (D) is superimposed on (E), showing similar trajectory, demonstrating that the vasculature structure identified in the tissue on the explanted array is likely of pia origin.

### 2.3 Length of implantation impacts the degree of material degradation and fibrous encapsulation

Based on the optical, TPM and SEM images, electrodes were assigned a binary score for the tip, shank, and degree of fibrous encapsulation (Figure 5). Electrodes that appeared to be broken or damaged by implantation/explantation were excluded from analysis. Tips and shanks were evaluated separately to examine the effects of both tip metallization and electrode insulation on device performance. The number of electrodes for each group are displayed in Table 1, excluding electrodes which were not wired or used for recording or stimulation. Differences in the total number of electrodes receiving a tip category (n=387), shank category (n=413), and encapsulation (n=380) are due to damage to the electrodes or excess encapsulation preventing the assignment of a proper category (Figures S1-3).

**Figure 5.**
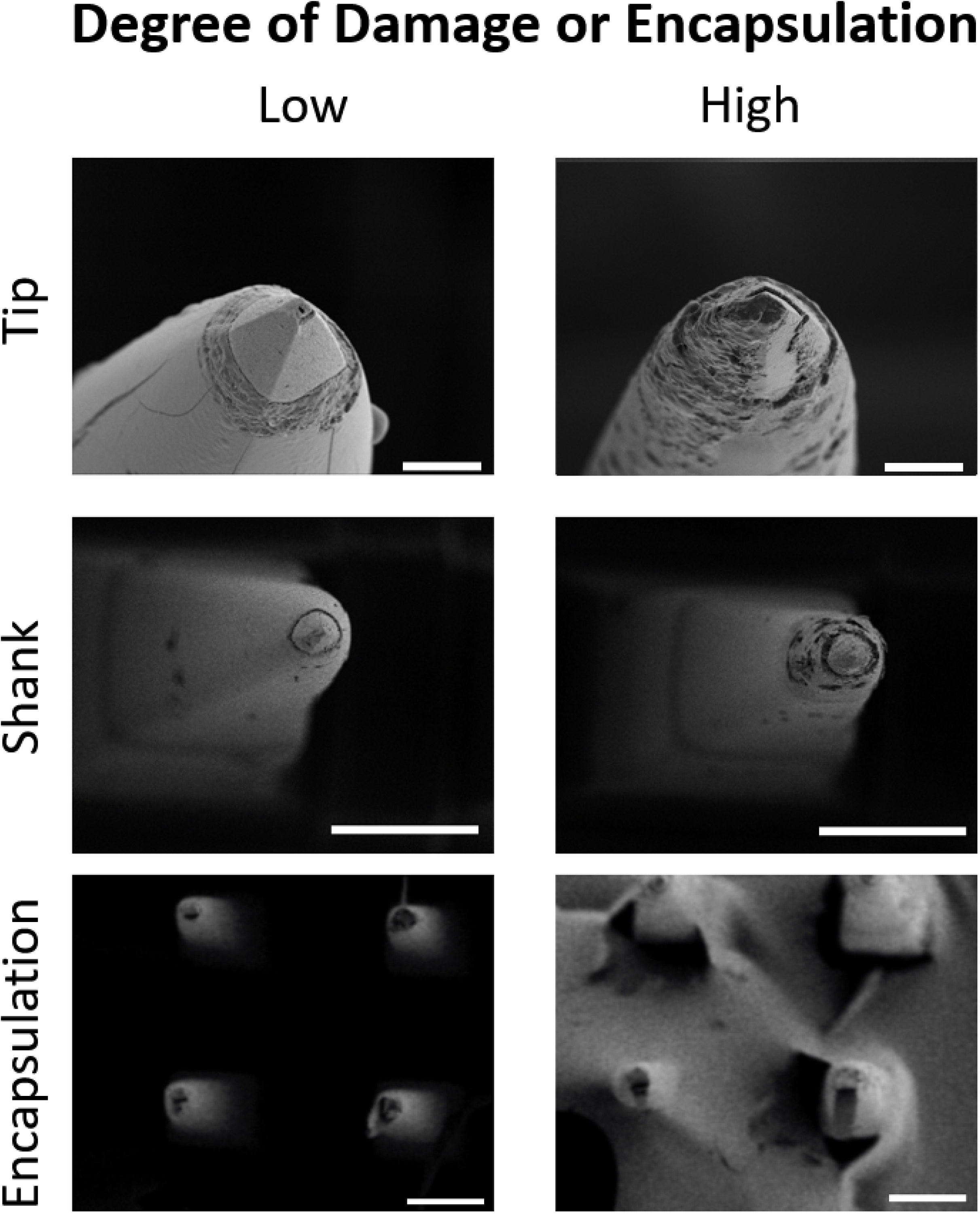
Tip and shank damage occurred on some implanted electrodes and encapsulation occurred on four implanted arrays. Representative high magnification images of undamaged/unencapsulated and damage/encapsulated electrodes. Tip images were taken from P1 array 1, with the degraded tip showing demetallation and biologic fouling (scale bar is 10µm). Shank images were taken from P2 lateral stimulating array (scale bare is 100µm). The degraded shank shows multiple surface and subsurface irregularities including pitting and delamination from the tip. Encapsulation images were from P2 medial stimulating array (scale bare is 100µm).

**Table 1.** Number of electrically connected electrodes that were classified as undamaged/unencapsulated or damaged/encapsulated based on tip degradation, shank degradation, and tissue encapsulation.

|  | Tip Degradation |  | Shank Degradation |  | Encapsulation |  |
| --- | --- | --- | --- | --- | --- | --- |
|  | Low (%) | High (%) | Low (%) | High (%) | Low (%) | High (%) |
| P1 | 122 (72.2) | 47 (27.8) | 151 (84.8) | 25 (15.2) | 51 (27.6) | 134 (72.4) |
| P2 Pt | 146 (90.1) | 16 (9.9) | 172 (98.3) | 3 (1.7) | 90 (50.6) | 88 (49.4) |
| P2 IrOx Medial | 26 (92.9) | 2 (7.1) | 32 (100) | 0 (0) | 6 (18.8) | 26 (81.2) |
| P2 IrOx Lateral | 7 (25) | 21 (75) | 9 (32.1) | 19 (67.9) | 30 (100) | 0 (0) |
| Total | 301 (77.7) | 86 (22.3) | 364 (88.8) | 46 (11.2) | 177 (41.6) | 248 (58.4) |
| Excluded | 45 |  | 22 |  | 7 |  |

Categories assigned to P1 and P2 platinum arrays were compared to identify any potential changes in material deterioration or encapsulation which may be attributed to the length of implantation (Table 2). We found that both measures of material degradation (tip and shank damage) were more prominent for longer implantation times (27.8% tip damage for P1 and 9.9% for P2, 15.2% shank damage for P1 and 1.7% for P2, p<0.001 for both). We also found that the degree of encapsulation is more significant for longer implants with 72.4% for the P1 arrays and 49.4% for the P2 arrays (p<0.001).

**Table 2.**
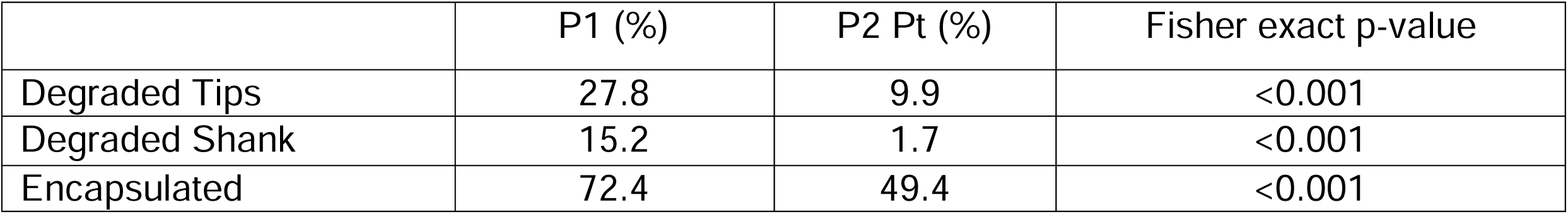
Differences observed between patients with different length of implant (980 days for P1 and 182 days for P2) on material degradation and encapsulation for electrically connected platinum recording electrode arrays

|  | P1 (%) | P2 Pt (%) | Fisher exact p-value |
| --- | --- | --- | --- |
| Degraded Tips | 27.8 | 9.9 | <0.001 |
| Degraded Shank | 15.2 | 1.7 | <0.001 |
| Encapsulated | 72.4 | 49.4 | <0.001 |

### 2.4 Stimulation resulted in electrode damage on one stimulating array but not the other

Two IrOx arrays implanted in P2 received a low amount of total charge (<160 µC per electrode site). Each of the two stimulated IrOx arrays had 60 electrodes, half of which electrically connected and used for stimulation. Preimplant optical images of the arrays did not show any variation between arrays. The stimulated electrode sites are arranged primarily in a checkerboard fashion. SEM shows that the lateral array had a high degree of tip and shank degradation (Figure 6A). Interestingly, tips and shanks showing visible damage appeared to coincide with the electrodes that were used for stimulation. Furthermore, EDS revealed that stimulated tips had lower iridium content than non-stimulated tips (Figure 6B). The loss of metallization for the lateral stimulating array occurred only on the electrodes used for stimulation. The medial array did not show this pattern (Figure 6C). The damage scores for each electrode tip and shank are summarized in the Figure 6D,E. The checkerboard pattern of damages of the lateral array is clearly seen, which correspond very well with arrangement of the stimulation electrodes. The medial stimulating array has overall much less observable material damage but more tissue encapsulation. Of the 62 electrodes used for stimulation on both arrays, 56 were analyzed, of which 23 had notable tip degradation, 21 of which were located on the lateral electrode array. Metal loss, and the corresponding decrease in iridium signal, was not observed on any non-stimulated electrodes. These results are summarized in Table 3.

**Figure 6.**
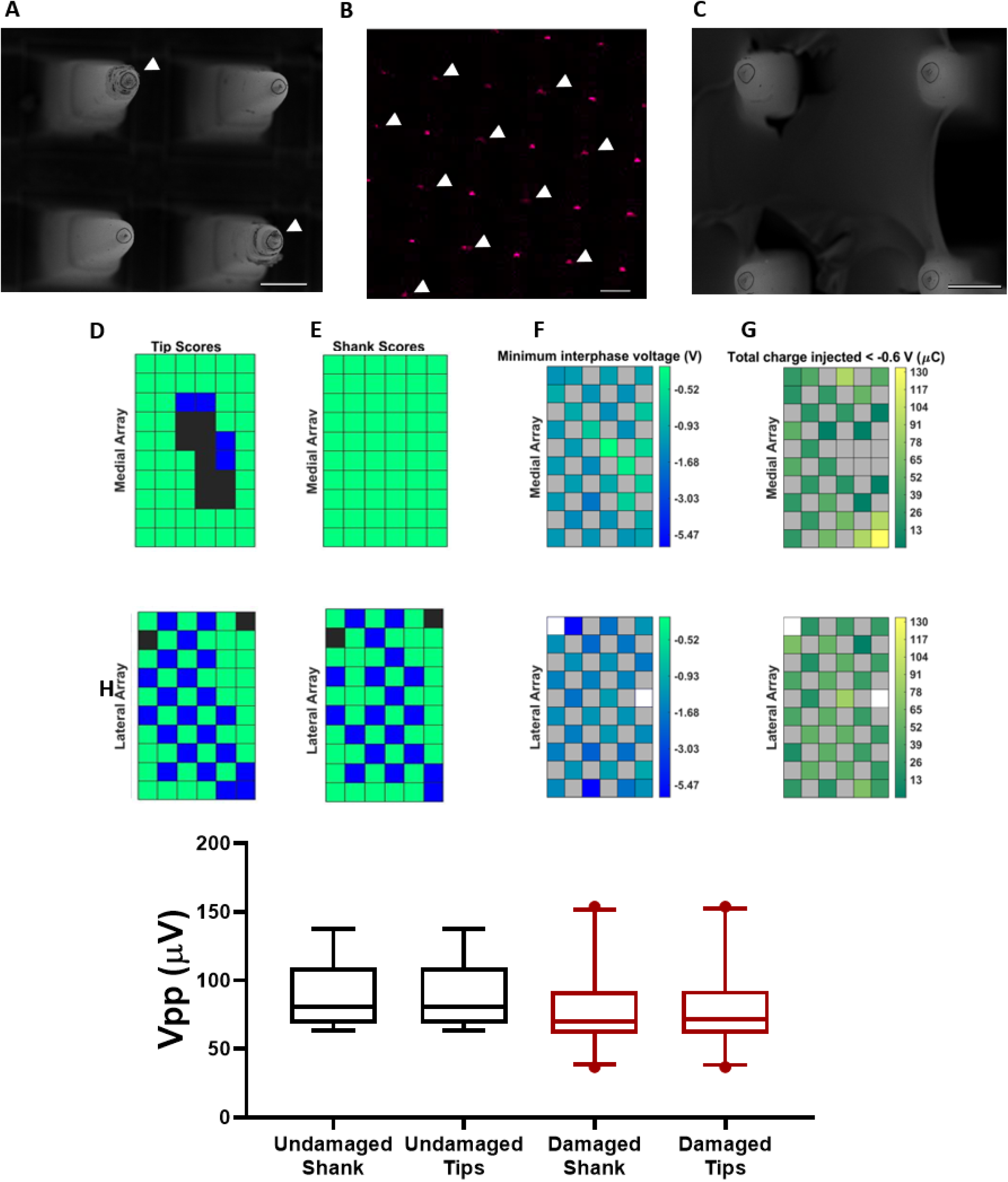
Stimulation-induced material damage on one of the two arrays. (A) SEM image of four shanks of the lateral array, tip damages are found on the stimulated electrodes marked with white arrows. (B) EDS of the stimulating electrodes for the lateral array showing reduced presence of iridium (magenta) on most of the stimulated sites (white arrows). (C) SEM image of the medial stimulating array tips. No differences were observed between the non-stimulated and stimulated tips on this array. Scale bars are 100µm. (D,E) Arrays showing the measured material properties on the stimulation arrays including tip categories (D) and shank categories (E). Green spaces show electrodes categorized as undamaged/unencapsulated, blue spaces show electrodes categorized as damaged, and black spaces show electrodes that were excluded from analysis. (F,G). Medial (top) and lateral (bottom) stimulating arrays showing (F) minimum interphase voltage (G) and total charge injected below -0.6V. The color bar for the minimum interphase voltages is log-transformed to emphasize differences between electrodes. Grey spaces indicate unwired electrodes. White spaces indicate wired electrodes that were never stimulated. (H) measured peak-to-peak voltages on stimulated electrodes after removing electrodes which were encapsulated with fibrous tissues. There were no significant differences observed in the measured unit amplitudes (Mann-Whitney non-parametric test)

**Table 3.** Effect of stimulation on material degradation and encapsulation for IrOx arrays. Non-stimulated tips were not electrically connected.

|  | Stimulated (%) | Non-stimulated (%) | X<sup>2</sup> Statistic | p-value |
| --- | --- | --- | --- | --- |
| Damaged Tips | 41.1 | 5.6 | 14.7 | <0.001 |
| Damaged Shank | 33.3 | 0.0 | 19.3 | <0.001 |
| High Encapsulation | 43.3 | 39.7 | 0.1 | >0.05 |

Delivered charge and measured interphase voltages were compared to the material degradation. The amount of stimulation provided was quantified by both the total charge delivered and number of pulses delivered. Although the mean amount of charge injected on the lateral array was greater, it was not significantly different than the mean charge injected on the medial array (Mann-Whitney test, p = 0.22). The medial array contained the three electrodes with the most charge delivered, none of which displayed observable material degradation. However, we examined the minimum voltage during the interphase period (Figure 6F) and found that the lateral array electrodes experienced higher voltage excursions on average than the medial array electrodes (mean minimum voltage was = -1.7 V for the lateral, and -1.1 V for the medial array). Furthermore, there was a significant relationship between the total charge injected at voltages more negative than -0.6V (Figure 6G) and the tip score (p = 0.025, crit-p = 0.034, logistic regression) and shank category (p = 0.023, crit-p = 0.034, logistic regression) on the lateral stimulating array. There was no relationship between total charge injected at voltages less than -0.6V and tip category (p = 0.60, logistic regression) or shank category (p = 1, logistic regression) on the medial array.

We found no significant differences in recording quality (Vpp) (Figure 6H) between the damaged and non-damaged electrodes on the two stimulation arrays, after excluding the encapsulated electrodes.

## 3. Discussion

In order for BCIs to become a viable therapy, the longevity of the devices and mechanisms of failure must be well understood. Effective electrode design requires knowledge of the stability of the materials in the harsh *in vivo* environment and the effects of gradually accumulating damage to the device. However, the relationships between chronic material degradation and device performance are poorly understood. The effects of material degradation on performance in human subjects is further complicated by the limited number of human subjects and the even smaller amount of explanted human BCI arrays. In this work, the *in vivo* performance of human neural electrode arrays was compared to the material integrity after explant. We have found signs of material degradation on all electrode arrays, with longer implantation times correlating with an increased number of degraded electrodes (Table 2). Additionally, biological tissue encapsulation on the explanted device was also documented as another potential factor to influence recording quality. The biological encapsulation tissues were highly collagenous and also highly cellularized, and appear to form in a time dependent manner, increasing with the length of implantation. A similar form of tissue response has been observed in a post mortem analysis of tissue surrounding a MEA implanted for seven months.(Szymanski et al. 2021) Further, the nature of the encapsulation at the periphery of the array and the center is different. Together, these results suggest that the encapsulation originated from the meninges, as opposed to the CNS.

### 3.1 Encapsulation and material degradation were both related to the length of implantation

Material and biological failure modes are most common on longer time scales (James et al. 2013), and it was expected that material degradation and collagenous encapsulation would increase with longer implant times. Indeed, we observed that the arrays implanted in P1 exhibited greater degrees of material degradation and encapsulation than those in P2 which were implanted for a much shorter length. We also observed a characteristic decline in recording performance and impedances with longer implantation times.

Impedance measurements are often used to determine the integrity of electrodes, while also serving as a method of investigating the interface between the electrode and the host tissues (Lago et al. 2016; Thakore et al. 2012). Previously, we have reported that in rats, complete fibrous encapsulation of the electrode resulted in lower 1kHz impedance compared to partial encapsulation (Cody et al. 2018). Complex impedance spectra analysis performed in the aforementioned study revealed unique features in the Nyquist plot that corresponds to an extracellular resistance component, which is smaller in the fully encapsulated device than the partially encapsulated device. This may be counterintuitive initially, but can be explained by a few mechanisms. First, the composition of the encapsulation tissue is high in collagen and less resistive than highly cellular and myelin rich brain tissue. Secondly, if the fibrotic growth at the base pushes the array up, a liquid filled gap will be formed between cone shaped shanks and the tract, creating a less resistive current path. Due to the limitation in our instrumentation in this study, impedance data were only obtained at 1 kHz preventing us from measuring complex impedances, but it is plausible that a similar effect may have occurred here. Full spectrum impedance recording in future studies could dissect the contributions from tissue encapsulation and material changes and determine whether the same factors are relevant here. However, such measurement needs to meet the regulatory requirements associated with clinical studies.

The decreases in impedance over time could also be a result of degradation of the electrode tips and shank insulation which leads to increased electrochemical surface area. Interestingly,we found no relationships between the impedance of the electrode at 1kHz and the Vpp during recording for Pt arrays (Figure S4). This is not surprising as impedance is only a measure of the electrochemical properties of the electrode and the electrode/tissue interface and does not account for biological variables such as distance from the electrode to the neuron or health of the host tissues, which are more relevant to Vpp. Impedance has previously been shown to be an unreliable predictor of recording performance in rodents and non-human primates (Cody et al. 2018; Jiang et al. 2014). Another important material factor that likely contribute to the uniform reduction of impedance on all arrays is the silicone hermetic sealing failure above the arrays from the wire bundle to the pedestal, which should be characterized in future studies.

The observed collagenous encapsulation of the arrays has been observed in rodent and non-human primate studies (Cody et al. 2018; Degenhart et al. 2016; James et al. 2013). Encapsulation of the electrode tip region can isolate the electrode from nearby neurons, resulting in a lowered Vpp. In addition, the collagenous material grown at the base of the array platform can lift the electrode up and away from the original target neurons, also resulting in Vpp decrease (Cody et al. 2018; Degenhart et al. 2016). Both tissue growth at the tips and the base have been observed from the explanted devices which may contribute to the degradation of Vpp over time in human subjects.

### 3.2 Stimulation at more negative voltages may drive material damage under certain circumstances

The stimulation parameters used in this study were based on studies from non-human primates showing that these parameters had no additional effects on cortical tissue apart from implanting the devices themselves, had no behavioral effect on the animal, and had limited effects on the electrode tissue interface (Chen et al. 2014; Kim et al. 2015). Here, we found that electrical stimulation induced damage on one of the two devices. On the lateral array, de-metallization was visible under SEM and detected by EDS only on the stimulated electrodes, indicating that stimulation was the cause of the metal loss. The reason that stimulation caused material damage on the lateral array but not the medial array is unclear. One notable difference between the lateral and medial array is that the medial had higher degree of tissue encapsulation. This can be a result of a higher degree of vascular damage or less stable fixation *in vivo*. While the specific reason for this cannot be determined, impedances were lower on the medial array (Figure S5). The decrease in impedance then could have resulted in lower amplitude voltage excursions during stimulation, decreasing the likelihood of material damage. Indeed, we found that the lateral array had more negative interphase voltages (mean = -1.7V) when compared to the medial array (mean = -1.1V).

More material damage was found on the lateral stimulating array in P2 which experienced higher cathodic interphase voltage amplitude. The interphase voltage is analogous to the maximum cathodic electrode potential (E_mc_) measured during charge injection limit (CIL) experiments performed *in vitro*. E_mc_ with values more negative than -0.6V (vs Ag/AgCl) are often considered to be unsafe due to irreversible water hydrolysis occurring at the electrode which could cause hydrogen gas production and pH increases (Cogan et al. 2005). Such reactions could lead to delamination of the IrOx coating even with a small number of pulses. We do not expect the median interphase voltage to be directly comparable to *in vitro* CIL measurements due to the two-electrode setup and increased variables introduced from the biological environment, but we expect the general relationship between voltage and interfacial reactions to hold. Besides the fact that lateral array experienced higher voltage excursion on average, we found a significant correlation between the charge injected below -0.6V and damage on the lateral stimulating array. These results indicate that stimulation, beyond a certain voltage threshold, may damage electrodes in a dose (charge) dependent manner.

Partial and complete loss of SIROF from Utah arrays upon continuous stimulation has been reported in previous in vitro studies (Negi et al. 2010), but the stimulation doses in these previous studies applied were much higher (7 h of continuous stimulation above 60nC). One potential explanation is that the stimulation on the lateral array only weakened the adhesion of the IrOx coating and the coating was stripped from the electrode during or after explantation. Alternatively, variations could also be a result of batch-to-batch difference in fabrication where the lateral array received poorly adhered IrOx coating. Notably, the electrodes damaged by stimulation performed just as well in recording as the undamaged electrodes. This surprising result indicates that despite the IrOx delamination and insulation cracking, the electrode is capable of recording neural signals (Hughes et al. 2020).

The biocompatibility of IrOx coatings has been widely studied and validated for stimulation and recording applications (Cogan 2008; Cogan et al. 2005; Hughes et al. 2020; Lee et al. 2002; Negi et al. 2010; Negi et al. 2010). In another of our studies, stimulating electrodes for over five years in a human participant did not result in worse signal recordings when compared to recording electrodes(Hughes et al. 2020). Furthermore, the ability to evoke sensation on stimulated electrodes only improved over time. Based on our observations here, this could be because a) the material damage caused by stimulation is idiosyncratic, and stimulation didn’t result in damage on the arrays of this 5-year study or b) material damage caused by stimulation had no effect on the electrode’s ability to record or stimulate. Discerning between the two is difficult, as studying the *in vivo* properties of the electrodes in parallel with the material properties is not possible in humans. Analysis will need to be conducted on these arrays that received much higher levels of stimulation after explant. Additionally, further animal studies using the stimulus parameters used in our study and assessing damage and changes in recording over time could provide insight here.

### 3.3 Implications for future intracortical electrode arrays

Overall, our results show that both material integrity and recording performance of human intracortical electrodes decrease over time. Degradation was observed on both electrode tip and the shank insulations. We have also observed different degree of tissue encapsulation both at the array base, middle of shank and tips of the arrays. Since we do not have real time data of these material and tissue changes, and explant analysis only provides a partial picture at the end point, we cannot accurately correlate material and biological factors to recording outcome. Multiple human studies have demonstrated that intracortical electrode recordings can enable brain-computer interface control of computer cursor and robotic arms for years after implant,(Bullard et al. 2020) yet the observations in the current study support the need for strategies for increasing material durability and decreasing fibrous encapsulation in order to further improve human BCI recording quality and longevity. Additionally, on one implanted array, we observed clear iridium loss as a result of stimulation, which correlated to more charge injected at more negative voltages. Further research on improving metal adhesion and developing real time electrode potential monitoring method during stimulation will eliminate such incidences.

## Data Availability

Data is available from corresponding author upon request

## Acknowledgements

We thank the participants for their dedication to this study. The recording and stimulation study was partly funded by the Defense Advanced Research Projects Agency’s (Arlington, VA, USA) Revolutionizing Prosthetics program (contract number N66001-10-C-4056). The views expressed herein are those of the authors and do not represent the official policy or position of the Department of Defense or US Government. The explant materials and tissue analysis were supported by the National Institute of Health grant R01NS110564, R01NS062109 and R01NS089688. We thank Dr. Pavel Takmakov of the Federal Drug and Agriculture for the initial discussion and cleaning protocols for SEM.

## Supplemental

**Supplemental Figure 1:**
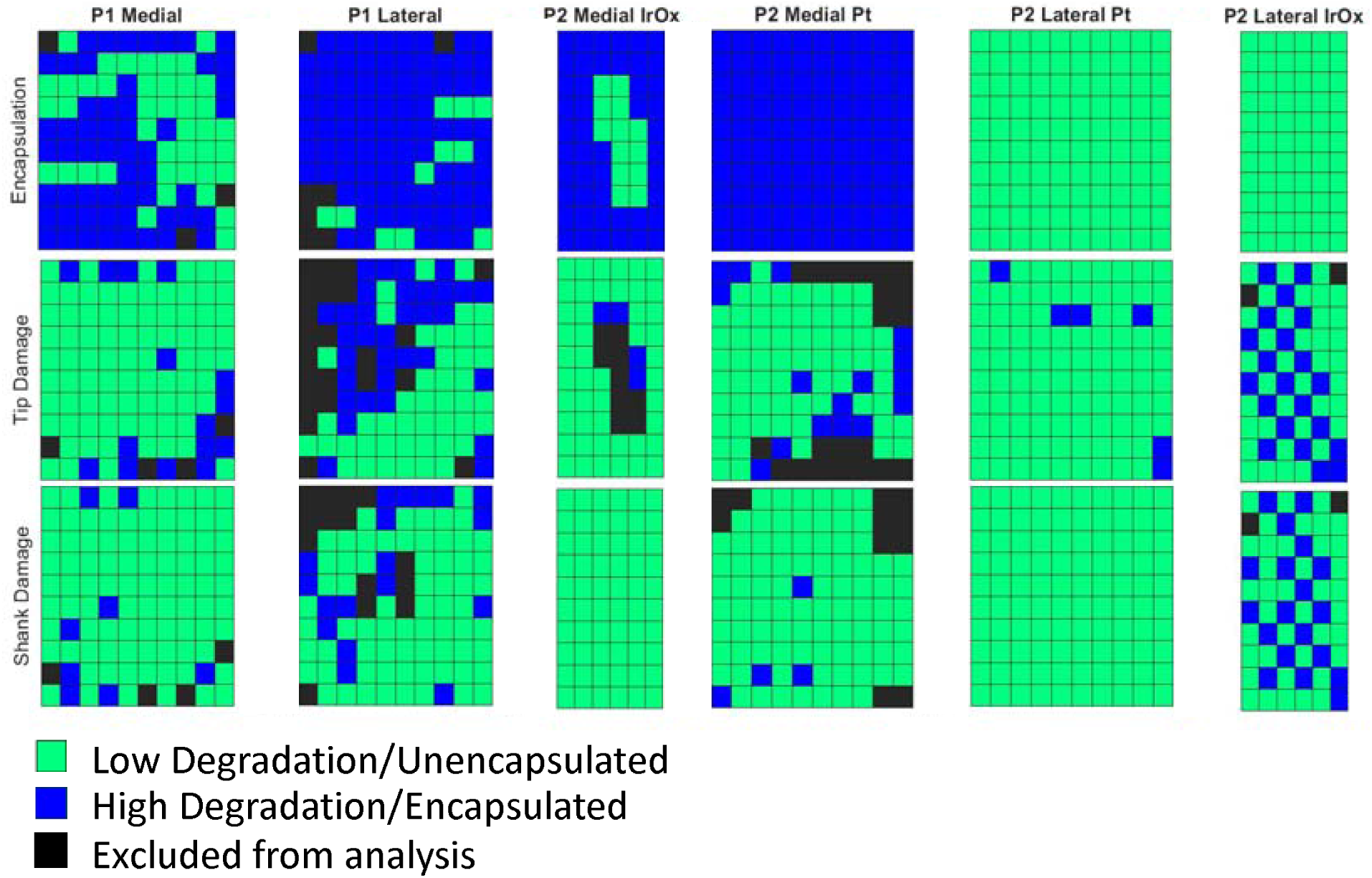
Categories assigned to each electrode site. Each site was assigned a category with respect to their material integrity or degree of encapsulation. Black sites were not able to be categorized and were excluded from the analysis.

**Supplemental Figure 2.**
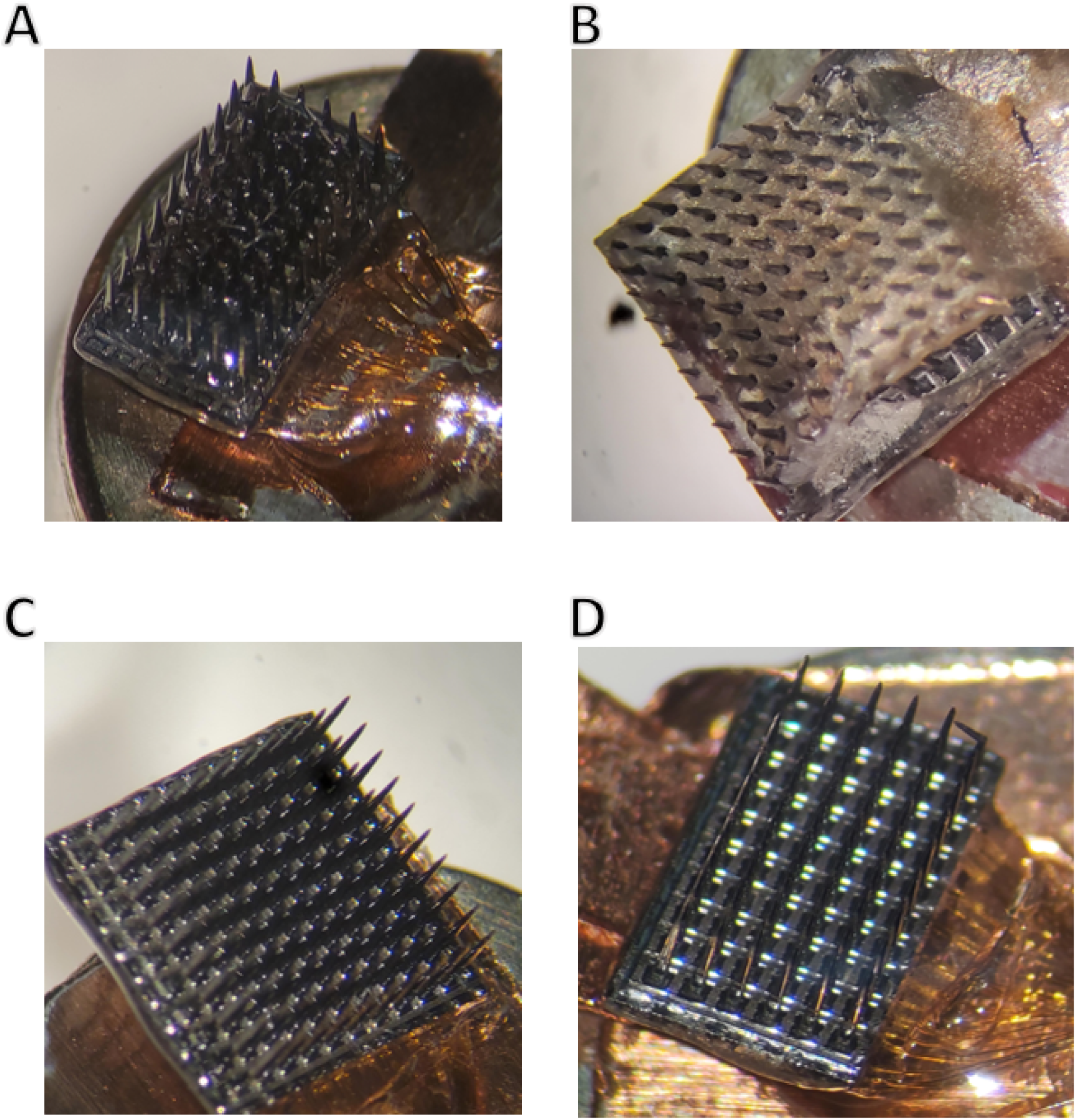
Optical images P2 arrays: A) Medial stimulating array from P2. B) Medial recording array from P2. C) Lateral recording array from P2. D) Lateral stimulating array from P2.

**Supplemental Figure 3.**
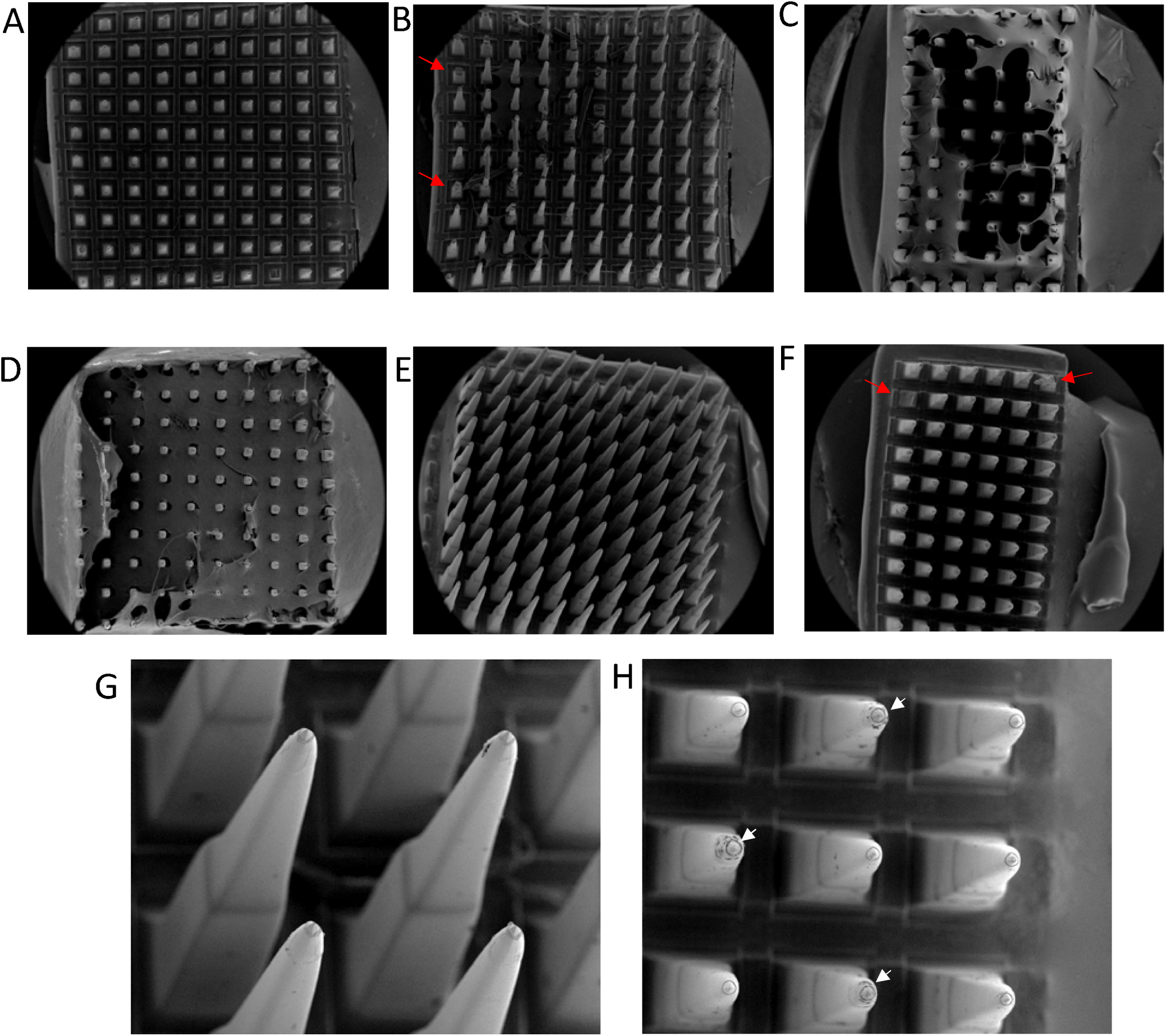
Scanning Electron Microscopy reveals damage and encapsulation on a fraction of implanted electrodes. A,B) Recording arrays implanted into P1 after enzymatic treatment. C) Medial stimulating array implanted into P2. D,E) medial and lateral recording arrays implanted into P2. F) lateral stimulating array implanted into P2. G,H) Higher magnification images of recording array in (E) and stimulating arrays in (F), respectively. White arrows indicate electrodes which were used for stimulation. Red arrows indicate representative electrodes which were excluded from analysis.

**Supplemental figure 4:**
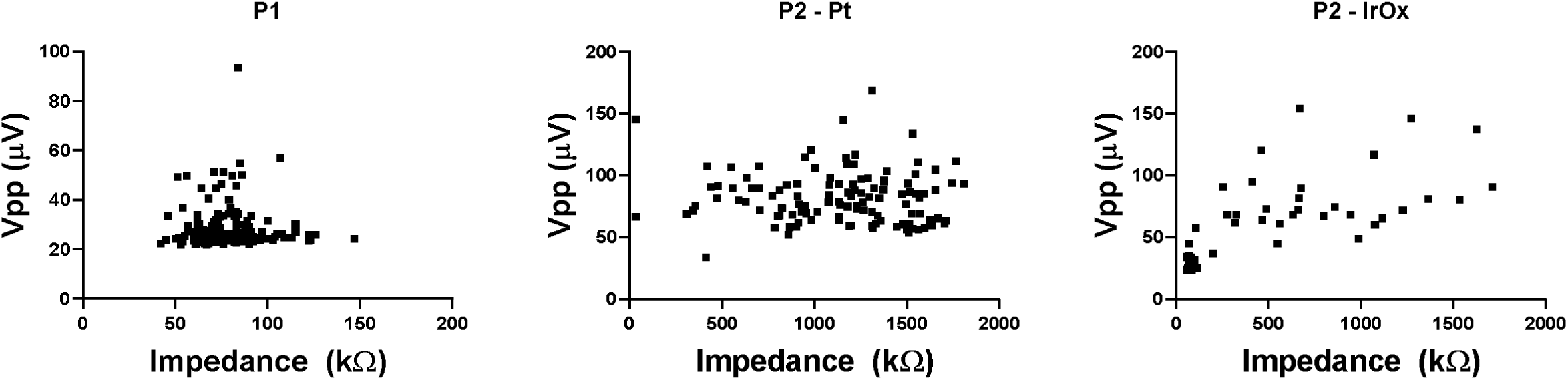
Comparisons of impedance and Vpp for P1, P2 platinum electrodes, and P2 IrOx electrodes on the last day of recording. No trends between impedance and Vpp were observed for either of the platinum tipped recording arrays.

**Supplemental Figure 5:**
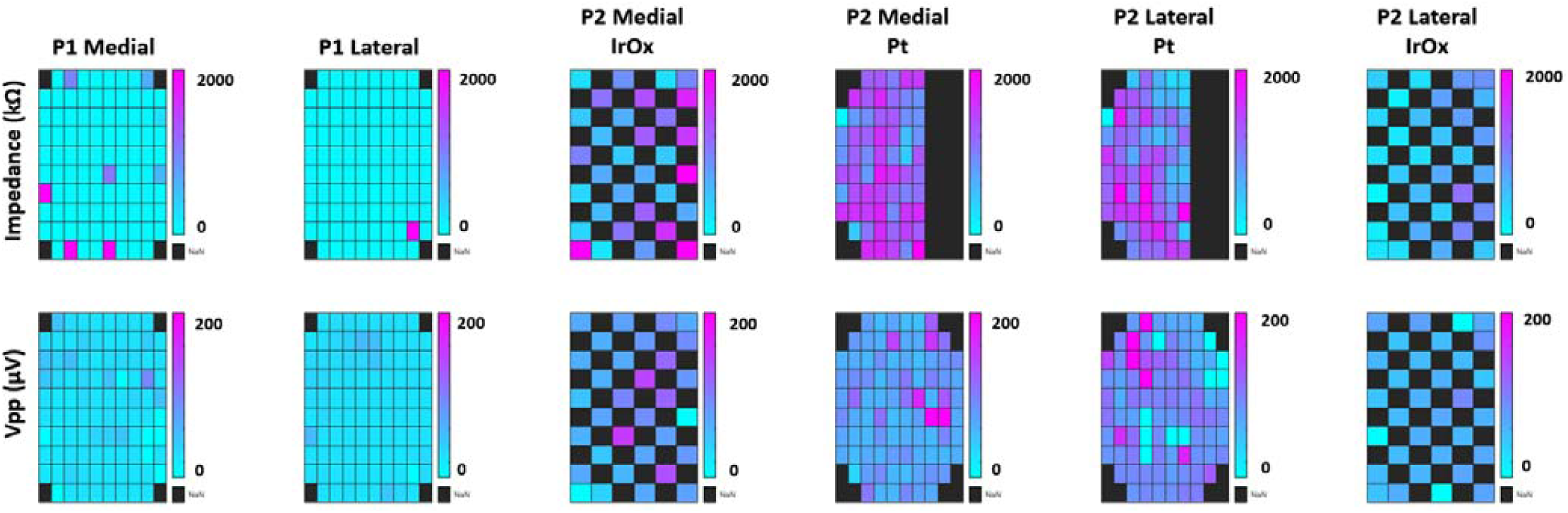
Impedances and Vpp for individual electrodes on the last day of recording. Lower impedances were observed on the arrays implanted in P1 due to the length of implantation. Impedances were not measured on every electrode in P2 due to hardware limitations, namely that the impedance cable could only measure impedances from a total of 96 channels across two arrays. Colors are linearly scaled.

